# Risk-Based Stratification Approach for Cardiovascular-Kidney-Metabolic Syndrome Stage 2 Using a Simple Algorithm

**DOI:** 10.64898/2026.01.28.26345070

**Authors:** Zhejia Tian, Maxim Sternberg, Jonas Willerding, Kai M. Schmidt-Ott, Anette Melk, Bernhard M. W. Schmidt

## Abstract

**Background:** Cardiovascular-kidney-metabolic (CKM) syndrome is staged to reflect increasing cardiometabolic risk. Substantial heterogeneity exists within stage 2, which includes individuals with metabolic risk factors or chronic kidney disease (CKD) without established cardiovascular disease and affect nearly half of US adults. This study aimed to develop and validate a sex-specific, clinically pragmatic model to subdivide CKM stage 2 into lower-risk (stage 2a) and higher-risk (stage 2b) subgroups

**Methods:** We analyzed participants aged ≥18 years with CKM syndrome stage 2 from the 1999-2018 NHANES cycles linked to the US National Death Index. Participants were assigned to development and validation cohorts comprising five survey cycles each. Predictor variables were selected using modified LASSO-regression and Cox-regression model, with cardiovascular mortality as the primary outcome. Model performance was assessed using time-dependent area under the receiver operating characteristic curve (AUC) and the C-statistics. TriNetX database with patient-level data from medical records was analyzed to provide clinical validation using major adverse cardiovascular events (MACE) as the outcome.

**Results:** Stage 2b in women was defined by the presence of at least two of the following: age ≥66 years, CKD, diabetes, or hypertension (AUC 0.79; C-index 0.78). In men stage 2b required at least two of the following: age ≥61 years, CKD, diabetes, or current smoking (AUC 0.73; C-index 0.72). Discrimination was preserved in external validation (women: AUC 0.70, C-index 0.69; men: AUC 0.75, C-index 0.68) with significantly distinct 10-year absolute risk difference of cardiovascular death (women: 4.9% [95% CI 2.2-7.5]; men: 8.5% [95% CI 5.0-12.0]). In TriNetX database, 10-year MACE risk increased from 11.3% to 24.6% in women and from 13.1% to 30.8% in men from stage 2a to stage 2b.

**Conclusions:** Subdividing CKM stage 2 into stages 2a and 2b identifies clinically meaningful differences in cardiovascular risk and may support targeted preventive strategies.

## Introduction

Given the strong interrelationships of metabolic syndrome (MetS), chronic kidney disease (CKD), and cardiovascular disease (CVD), the American Heart Association (AHA) conceptualized in 2023 the cardiovascular-kidney-metabolic (CKM) syndrome to promote integrated CKM health in the population^1,2^. The CKM syndrome describes a progressive condition and has been classified into five stages, ranging from absence of risk factors (stage 0) through escalating cardiovascular risk without established CVD (stage 1 and 2) to advanced stages with subclinical and clinical CVD (stage 3 and 4) (Supplemental Table 1)^1,2^.

The CKM syndrome is associated with a graded increase in cardiovascular morbidity and mortality across the five stages^3^. Using the National Health and Nutrition Examination Survey (NHANES) data, we and others estimated the prevalence of each CKM syndrome stage, and individuals classified as stage⍰2 comprise almost half of the US adult population with notable age- and sex-disparities^4–8^. CKM syndrome stage 2 includes individuals with metabolic risk factors and/or moderate-to high-risk CKD without CVD. For patients in stage 2, the primary emphasis is on lifestyle modification and pharmacologic treatment of modifiable risk factors to prevent the onset of CVD and progression of CKD leading to kidney failure. However, achieving these goals is challenging given that this subgroup accounts for roughly half of the adult population. Thus, precise patient care requires better identification of high⍰risk individuals within this subgroup.

Using NHANES data, we aim to develop a sex-specific risk-stratification model that subdivides CKM⍰syndrome stage⍰2 into two risk groups based on easily assessable parameters while retaining acceptable discrimination accuracy. The model is further validated in an independent cohort drawn from the TriNetX database, confirming its discriminative performance.

## Materials and Methods

### Development and Validation Cohorts

NHANES is a biannual, cross-sectional survey conducted since 1999, utilizing a complex, multistage, probability sampling design to represent the non-institutionalized civilian US population. Written informed content was obtained from all survey participants, and the study procedures receive approval from the National Center for Health Statistics Research Ethics Review Board. This study received a waiver from the institutional review board at Hannover Medical School, since only de-identified patient data were included.

Data were derived from the NHANES 1999-2018 cycles, linked to the U.S. National Death Index. NHANES 1999-2000, 2003-2004, 2007-2008, 2011-2012, 2015-2016 cycles were merged to form the model development cohort, while the remaining cycles were used for validation (validation cohort). Non-pregnant adults aged ≥18 years with sufficient information to ascertain CVD status based on self-report were included. Participants were required to have completed both the physical examination and laboratory components of the survey with available examination weights.

### Data Collection

Participants in CKM syndrome stage 2 were identified consistent with the definitions^1^ using demographic, medication, and health behavior data obtained from household interviews, as well as clinical examination findings and laboratory measurements.

Hypertension was defined by elevated blood pressure meeting 2025 AHA guideline^9^ criteria or by the use of antihypertensive medication. Blood pressure values were derived from the mean of the last two blood pressure measurements^10^. Moderate-to high-risk CKD was classified in accordance with the 2024 KDIGO guidelines^11^. Estimated glomerular filtration (eGFR) was calculated with the 2021 race- and ethnicity-free Chronic Kidney Disease Epidemiology Collaboration creatinine equation^12^. Urinary albumin to creatinine ratio (UACR) was calculated from urinary albumin and urinary creatinine. Normal lipid condition was determined based on 2018 AHA guideline recommendations^13^. Diabetes status was assessed using self-reported antidiabetic therapy, glycated hemoglobin, and fasting serum glucose levels. The presence of diabetes or prediabetes was determined according to diagnostic criteria outlined in the 2025 ADA guidelines^14^. 10-year CVD risk was estimated with PREVENT equations^15,16^. Values for age, blood pressure and lipids outside the definition of the PREVENT score were adjusted to the respective upper or lower allowed extreme value.

### Clinical Validation Using Real-World Data

We applied the model to TriNetX database to evaluate its performance in a real-world clinical setting. TriNetX US Collaborative Network provides access to de-identified electronic medical records from approximately 126 million patients, both inpatients and outpatients, across 69 healthcare organizations. Electronic medical records include sociodemographic data; diagnoses coded using the International Classification of Diseases, Tenth Revision, Clinical Modification (ICD-10-CM); procedures recorded with either the ICD-10 Procedure Coding System (ICD-10-PCS) or Current Procedural Terminology (CPT); medications coded according to the Veterans Affairs National Formulary; and laboratory tests classified using Logical Observation Identifiers Names and Codes (LOINC)^17^.

In TriNetX database, we included adults aged between 18 and 90 years who fulfilled the criteria for CKM syndrome stage 2 (Supplemental Table S2-5). Pregnant participants during the follow-up were excluded.

### Outcome Ascertainment

For NHANES cohort, the primary outcome was cardiovascular death over ten years. For clinical cohort, the primary outcome was major adverse cardiac events (MACE) during 10-year follow-up, as cause-specific mortality data was not available in TriNetX database. MACE was defined as a composite of the following events with associated ICD-10-CM codes: fatal or nonfatal acute or subsequent myocardial infarction in any territory (I21-22), fatal or nonfatal cardiac arrest (I46), fatal or nonfatal cerebral infarction in any territory (I63), fatal or nonfatal nontraumatic intracerebral hemorrhage (I61), and all-cause mortality. Mortality data were derived from death records, billable codes from closed claims, Social Security Administration Master Death File, private obituaries, and private claims.

### Statistical Analysis

For analysis of the NHANES database, we accounted for complex survey design following the National Center for Health Statistic analytic guideline^18^. Baseline characteristics were presented as means ± SE for continuous variables and as frequency or proportion for categorial variables. For variables with skewed distributions, natural logarithmic transformation was applied as appropriate. Missing values were addressed with multivariate multiple imputation by chained equations^19^.

For model development, an adjusted least absolute shrinkage and selection operator (LASSO) regression was first applied for preliminary variable selection, incorporating sampling weights in the estimation process^20^. Variables that minimized error based on the bootstrap replicating weights method were selected, and CKM syndrome stage 2 conditions related to these variables were subsequently included in multivariable Cox proportional hazards models for final selection. Model improvement was assessed using design-based Wald test (F-statistics)^21^. The marginal effect of age on the predicted hazard of cardiovascular death was analyzed, and the age at which the risk began to rise steeply was identified. The final model was determined using the maximally selected Wald statistic^22^ with the largest z-statistic to distinguish high⍰risk from low⍰risk individuals. Internal validation of the predictive model was conducted only within the development cohort using a bootstrap resampling approach to assess and correct for overfitting^23^. The discrimination was evaluated with concordance statistics (C-statistic) and weighted time-dependent area under the receiver operating characteristic curve (AUC) based on bootstrap approach^24,25^. For latter, bootstrap replicate weights were first generated according to strata and primary sampling units of NHANES complex survey design. Then, for each bootstrap sample, the model was re-estimated, and the time-dependent AUC was recalculated. Finally, the average AUC and its 95% confidence interval were obtained from all bootstrap samples. Model calibration was assessed by comparing predicted risks from the Cox proportional hazards model with observed risks. Survival probabilities were estimated using survey-weighted Kaplan-Meier methods in NHANES database^21^. Absolute risk difference between CKM syndrome stage 2a and stage 2b were calculated at the end of 10-year follow-up.

For analysis using TriNetX database, missing data of covariates were imputed by TriNetX build-in statistical tool. Participants were censored the day after their last clinical record within the follow-up period. Survival analysis was performed using Kaplan-Meier estimate. Differences in primary outcome by stage status and sex was analyzed using a Cox proportional hazards regression model and was presented as hazard ratios (HRs) and 95% CIs.

All tests were 2-sided, and P < 0.05 was considered statistically significant for hypothesis testing. Statistical analyses were conducted using R version 4.3.2. or the built-in statistical tools of the TriNetX research platform (Java 11.0.16; R 4.0.2; Python 3.7).

## Results

### Baseline Characteristics

The development cohort comprised 2,596 female and 2,664 male adults in CKM syndrome stage 2, corresponding to 11,333,846 noninstitutionalized US women and 12,253,156 men (Table 1). Compared with men, the women were older and slightly more likely to belong to racial/ethnic minority groups. They exhibited higher prevalence of MetS, CKD, and use of antihypertensive medication, but lower prevalence of hypertriglyceridemia and nicotine dependence. The mean PREVENT score for women was higher than that for men. In the validation cohort, 2,757 female and 2,772 male adults meeting criteria for CKM⍰syndrome stage⍰2 were identified. When weighted to the US population, these participants were representative of 11,631,674 women and 12,283,114 men, respectively. By sex, the baseline characteristics of the validation cohort were comparable to those of the development cohort (Table 1).

**Table 1.**
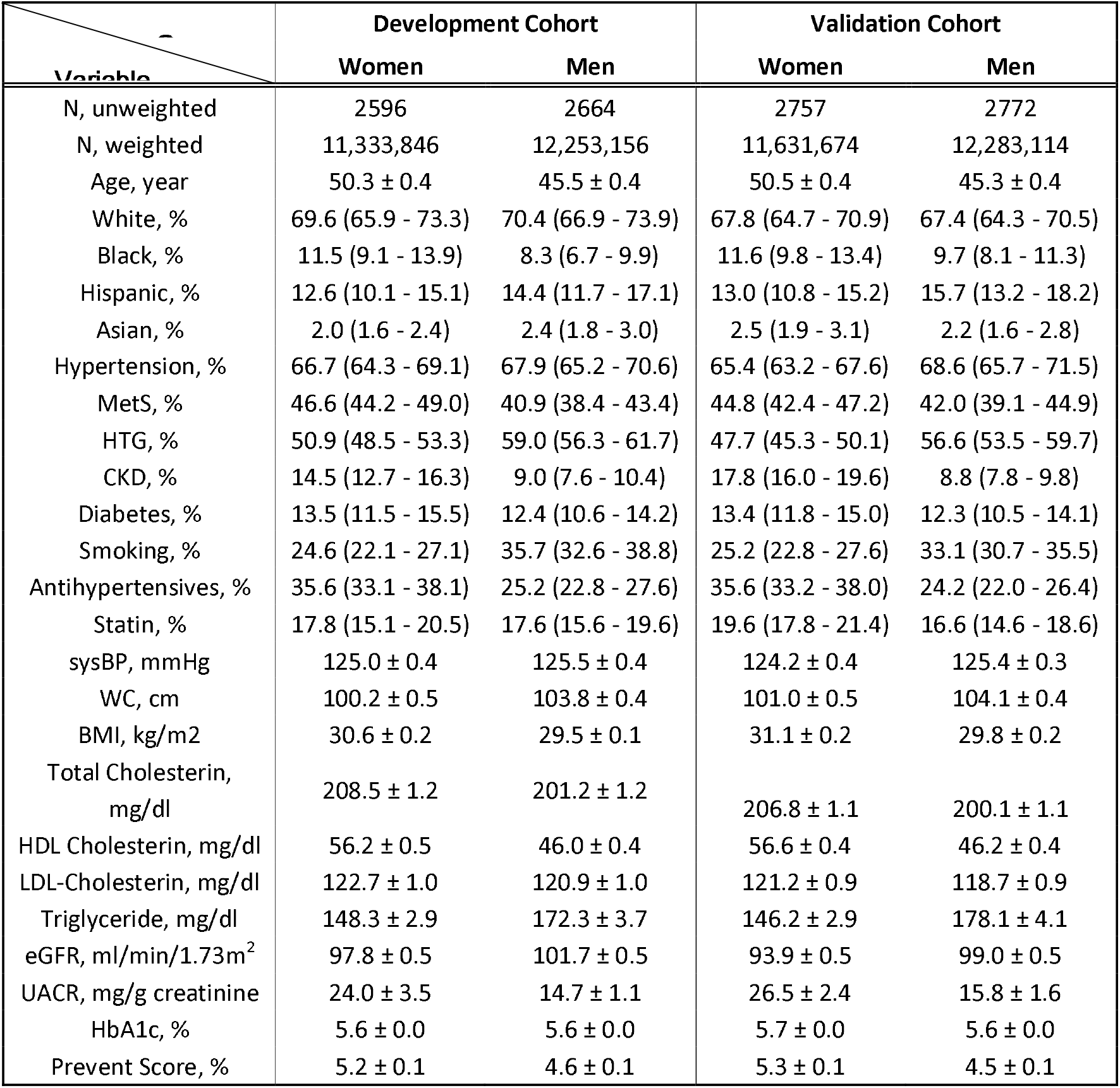
Baseline Characteristics of Development and Validation Cohorts. Means are presented as weighted estimates with standard error. Prevalences are presented as weighted estimates with 95% confidence intervals. Abbreviations: BMI, body mass index; CKD, chronic kidney disease; eGFR, estimated glomerular filtration rate; HbA1c, glycated hemoglobin; HDL, high density lipoprotein; HTG, hypertriglyceridemia; LDL, low density lipoprotein; MetS, metabolic syndrome; sysBP, systolic blood pressure; UACR, urine albumin creatinine ratio, WC, waist circumference.

| Variable | Development Cohort |  | Validation Cohort |  |
| --- | --- | --- | --- | --- |
|  | Women | Men | Women | Men |
| N, unweighted | 2596 | 2664 | 2757 | 2772 |
| N, weighted | 11,333,846 | 12,253,156 | 11,631,674 | 12,283,114 |
| Age, year | 50.3 ± 0.4 | 45.5 ± 0.4 | 50.5 ± 0.4 | 45.3 ± 0.4 |
| White, % | 69.6 (65.9 - 73.3) | 70.4 (66.9 - 73.9) | 67.8 (64.7 - 70.9) | 67.4 (64.3 - 70.5) |
| Black, % | 11.5 (9.1 - 13.9) | 8.3 (6.7 - 9.9) | 11.6 (9.8 - 13.4) | 9.7 (8.1 - 11.3) |
| Hispanic, % | 12.6 (10.1 - 15.1) | 14.4 (11.7 - 17.1) | 13.0 (10.8 - 15.2) | 15.7 (13.2 - 18.2) |
| Asian, % | 2.0 (1.6 - 2.4) | 2.4 (1.8 - 3.0) | 2.5 (1.9 - 3.1) | 2.2 (1.6 - 2.8) |
| Hypertension, % | 66.7 (64.3 - 69.1) | 67.9 (65.2 - 70.6) | 65.4 (63.2 - 67.6) | 68.6 (65.7 - 71.5) |
| MetS, % | 46.6 (44.2 - 49.0) | 40.9 (38.4 - 43.4) | 44.8 (42.4 - 47.2) | 42.0 (39.1 - 44.9) |
| HTG, % | 50.9 (48.5 - 53.3) | 59.0 (56.3 - 61.7) | 47.7 (45.3 - 50.1) | 56.6 (53.5 - 59.7) |
| CKD, % | 14.5 (12.7 - 16.3) | 9.0 (7.6 - 10.4) | 17.8 (16.0 - 19.6) | 8.8 (7.8 - 9.8) |
| Diabetes, % | 13.5 (11.5 - 15.5) | 12.4 (10.6 - 14.2) | 13.4 (11.8 - 15.0) | 12.3 (10.5 - 14.1) |
| Smoking, % | 24.6 (22.1 - 27.1) | 35.7 (32.6 - 38.8) | 25.2 (22.8 - 27.6) | 33.1 (30.7 - 35.5) |
| Antihypertensives, % | 35.6 (33.1 - 38.1) | 25.2 (22.8 - 27.6) | 35.6 (33.2 - 38.0) | 24.2 (22.0 - 26.4) |
| Statin, % | 17.8 (15.1 - 20.5) | 17.6 (15.6 - 19.6) | 19.6 (17.8 - 21.4) | 16.6 (14.6 - 18.6) |
| sysBP, mmHg | 125.0 ± 0.4 | 125.5 ± 0.4 | 124.2 ± 0.4 | 125.4 ± 0.3 |
| WC, cm | 100.2 ± 0.5 | 103.8 ± 0.4 | 101.0 ± 0.5 | 104.1 ± 0.4 |
| BMI, kg/m<sup>2</sup> | 30.6 ± 0.2 | 29.5 ± 0.1 | 31.1 ± 0.2 | 29.8 ± 0.2 |
| Total Cholesterol, mg/dl | 208.5 ± 1.2 | 201.2 ± 1.2 | 206.8 ± 1.1 | 200.1 ± 1.1 |
| HDL Cholesterol, mg/dl | 56.2 ± 0.5 | 46.0 ± 0.4 | 56.6 ± 0.4 | 46.2 ± 0.4 |
| LDL-Cholesterol, mg/dl | 122.7 ± 1.0 | 120.9 ± 1.0 | 121.2 ± 0.9 | 118.7 ± 0.9 |
| Triglyceride, mg/dl | 148.3 ± 2.9 | 172.3 ± 3.7 | 146.2 ± 2.9 | 178.1 ± 4.1 |
| eGFR, ml/min/1.73m<sup>2</sup> | 97.8 ± 0.5 | 101.7 ± 0.5 | 93.9 ± 0.5 | 99.0 ± 0.5 |
| UACR, mg/g creatinine | 24.0 ± 3.5 | 14.7 ± 1.1 | 26.5 ± 2.4 | 15.8 ± 1.6 |
| HbA1c, % | 5.6 ± 0.0 | 5.6 ± 0.0 | 5.7 ± 0.0 | 5.6 ± 0.0 |
| Prevent Score, % | 5.2 ± 0.1 | 4.6 ± 0.1 | 5.3 ± 0.1 | 4.5 ± 0.1 |

### Derivation of the Risk-Stratification Model in the Development Cohort

Adjusted LASSO regression first selected eight of the 13 candidate variables each for women and for men, using the λ that minimizes the average prediction error as the tuning parameter (logλ_min_ = ×6.87 for women; logλ_min_ = ×6.52 for men) (Supplemental Figure S1). For women, age⍰66 years was identified as the point at which the risk began to increase steeply, whereas for men the corresponding threshold was age⍰61⍰years (Supplemental Figure S2). The conditions associated with the selected variables were then entered sequentially into multivariable Cox⍰regression models, as summarized in Supplemental Table S6. The final set of variables retained in the model was, for women: age⍰≥⍰66⍰years, CKD, diabetes, and hypertension; and for men: age⍰≥⍰61⍰years, CKD, diabetes, and smoking. Using the maximally selected Wald statistic, the optimal threshold for defining stage⍰2b was identified as the presence of at least two of the four selected variables (largest⍰|z|⍰=⍰8.65 for women; largest⍰|z|⍰=⍰5.57 for men). Individuals meeting this criterion were classified as CKM syndrome stage⍰2b, whereas those with fewer than two variables were assigned to stage⍰2a.

### Performance Assessment in the Development Cohort

In women, the risk-stratification model achieved a median C⍰statistic of 0.78 (95⍰%⍰CI⍰0.75-0.83) and a time⍰dependent weighted AUC of 0.79 (95⍰%⍰CI⍰0.74-0.85) (Figure 1A). In men, the corresponding median C⍰statistic was 0.72 (95⍰%⍰CI⍰0.68-0.76) and the time⍰dependent weighted AUC was 0.73 (95⍰%⍰CI⍰0.68-0.78) (Figure 1B). Among women in the development samples, the calibration slope median was 0.92 (interquartile range [IQI] 0.91-0.93) (Figure 1C), whereas among men, it was 0.87 (IQI 0.86-0.89) (Figure 1D). The internal validation of the model was displayed in the density plot (Supplemental Figure S3 and S4). The HR obtained from the original Cox regression model was comparable to the mean HR derived from the bootstrap resampling, indicating good internal reproducibility. After implementing the risk stratification model, 74.9⍰% of US adults (33.3⍰% women vs. 41.6⍰% men) in the development cohort fell into low⍰risk stage⍰2a, while 25.1⍰% (15.9⍰% women vs. 9.2⍰% men) were assigned to high⍰risk stage⍰2b (Figure 2A).

**Figure 1.**
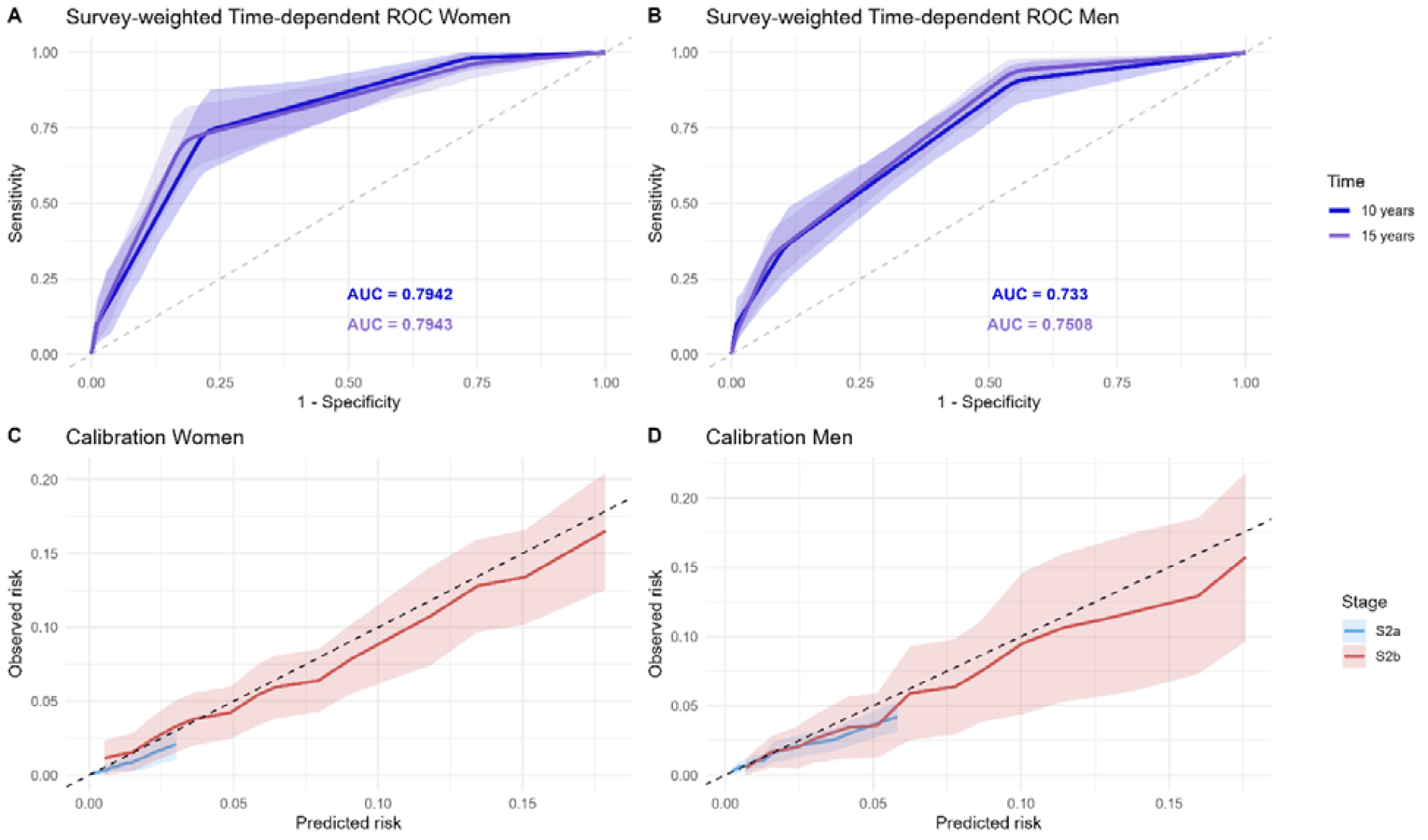
Internal Performance Assessments of the Sex-specific Risk-Stratification Model. Panels A and B present time-dependent ROC curves illustrating the discriminative performance of the sex-specific risk-stratification model for cardiovascular death in women (A) and men (B). Panels C and D show calibration plots for the same model in women (C) and men (D). *Abbreviations: AUC, area under the receiver operating characteristic curves; ROC, receiver operating characteristic*.

**Figure 2.**
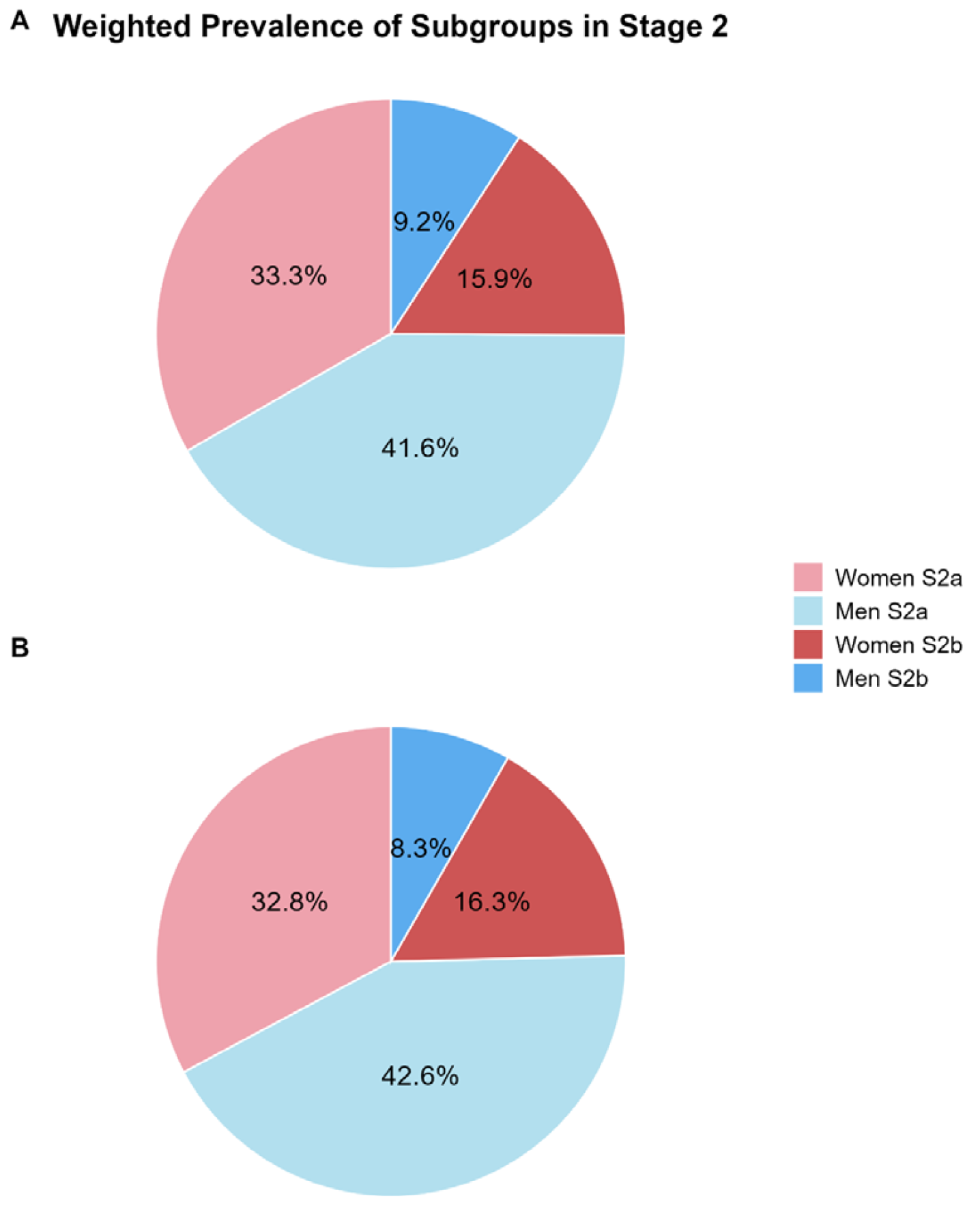
Sex-specific Prevalence of CKM syndrome Stage 2a and Stage 2b within Stage 2. Panels A and B depict the weighted prevalence of CKM syndrome stage 2a and 2b among individuals with CKM syndrome stage 2 in the development cohort (A) and validation cohort (B). *Abbreviations: CKM, cardiovascular-kidney-metabolic*.

The 10-year cumulative incidence of cardiovascular death among women in CKM stage 2a was 0.8% (95% CI 0.3-1.4) compared with 6.4% (95% CI 4.3-8.5) among those in stage 2b, yielding an absolute risk difference of 5.6% (95% CI 3.4-7.8) (Figure 3A). Among Men, the corresponding incidences were 2.4% (95% CI 1.6-3.2) for stage 2a and 9.5% (95% CI 4.4-14.6) for stage 2b, with an absolute risk difference of 7.1% (95% CI 1.9-12.3) (Figure 3B).

**Figure 3.**
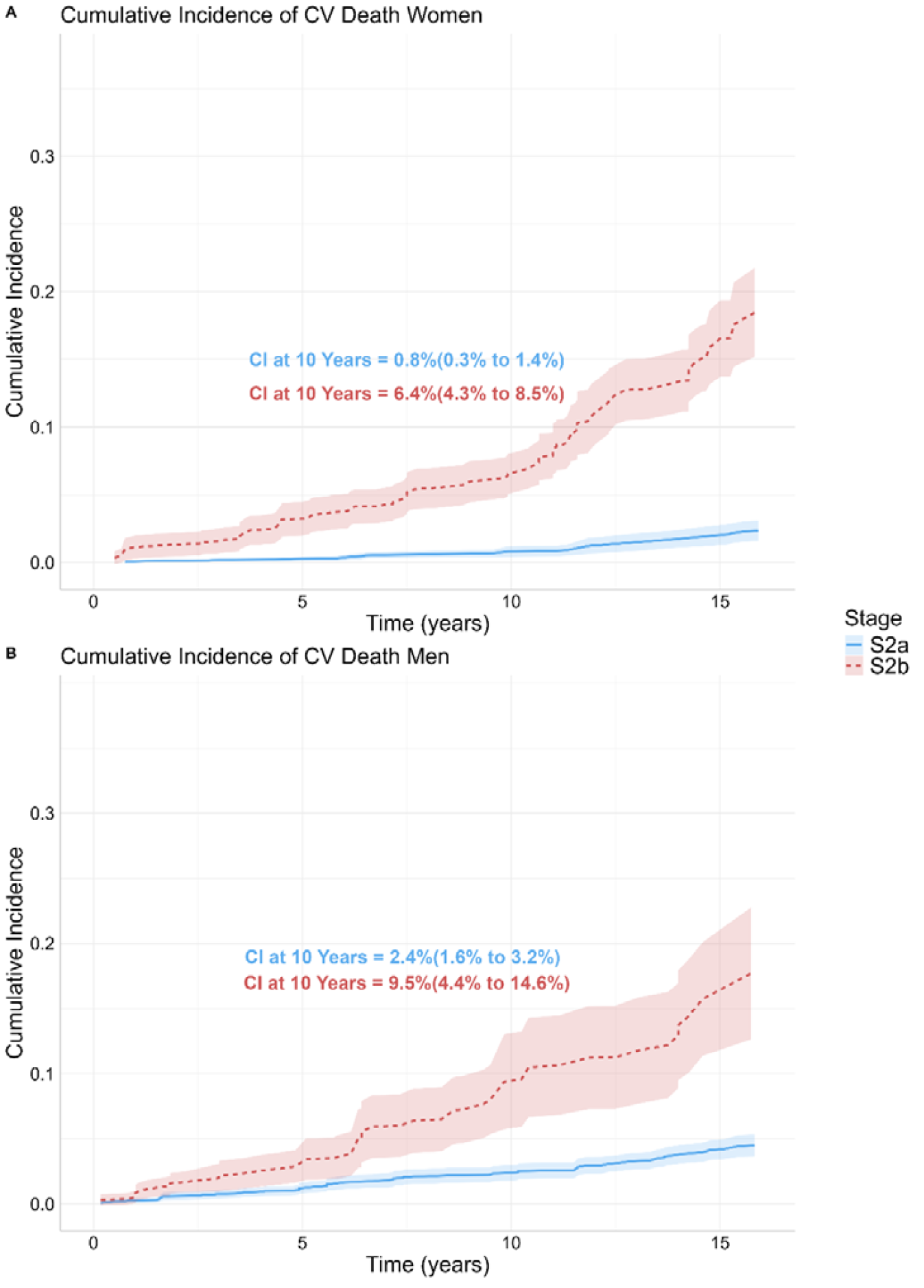
10-Year Cumulative Incidence of Cardiovascular Death among Participants with CKM Syndrome Stage 2a and Stage 2b in the Development Cohort. Panels A and B depict weighted cumulative incidence curves for cardiovascular death in women and men, respectively, stratified by CKM syndrome stage 2a and 2b, with 95% confidence intervals. 10-year cumulative incidences are additionally presented as point estimates with 95% confidence intervals. *Abbreviations: CKM, cardiovascular-kidney-metabolic; CI, cumulative incidence*.

### External Validation

In the validation cohort, 75.4% of US adults (32.8⍰% women vs. 42.6⍰% men) were assigned to CKM syndrome stage 2a, yet the remaining participants (16.3⍰% women vs. 8.3⍰% men) met the criterion for stage 2b (Figure 2B). This risk-stratification model among women demonstrated a median C statistic of 0.69 (95% CI 0.63-0.75) and a time-dependent weighted AUC of 0.70 (95% CI 0.62-0.77) (Figure 4A). Among men, the median C statistic was 0.68 (95% CI 0.60 - 0.76) and the time-dependent weighted AUC was 0.75 (95% 0.69-0.80) (Figure 4B). Calibration plots are presented in Figure 4C-D. The median calibration slope was 0.96 (IQI 0.95-0.96) for women and 1.04 (IQI 1.02-1.06) for men.

**Figure 4.**
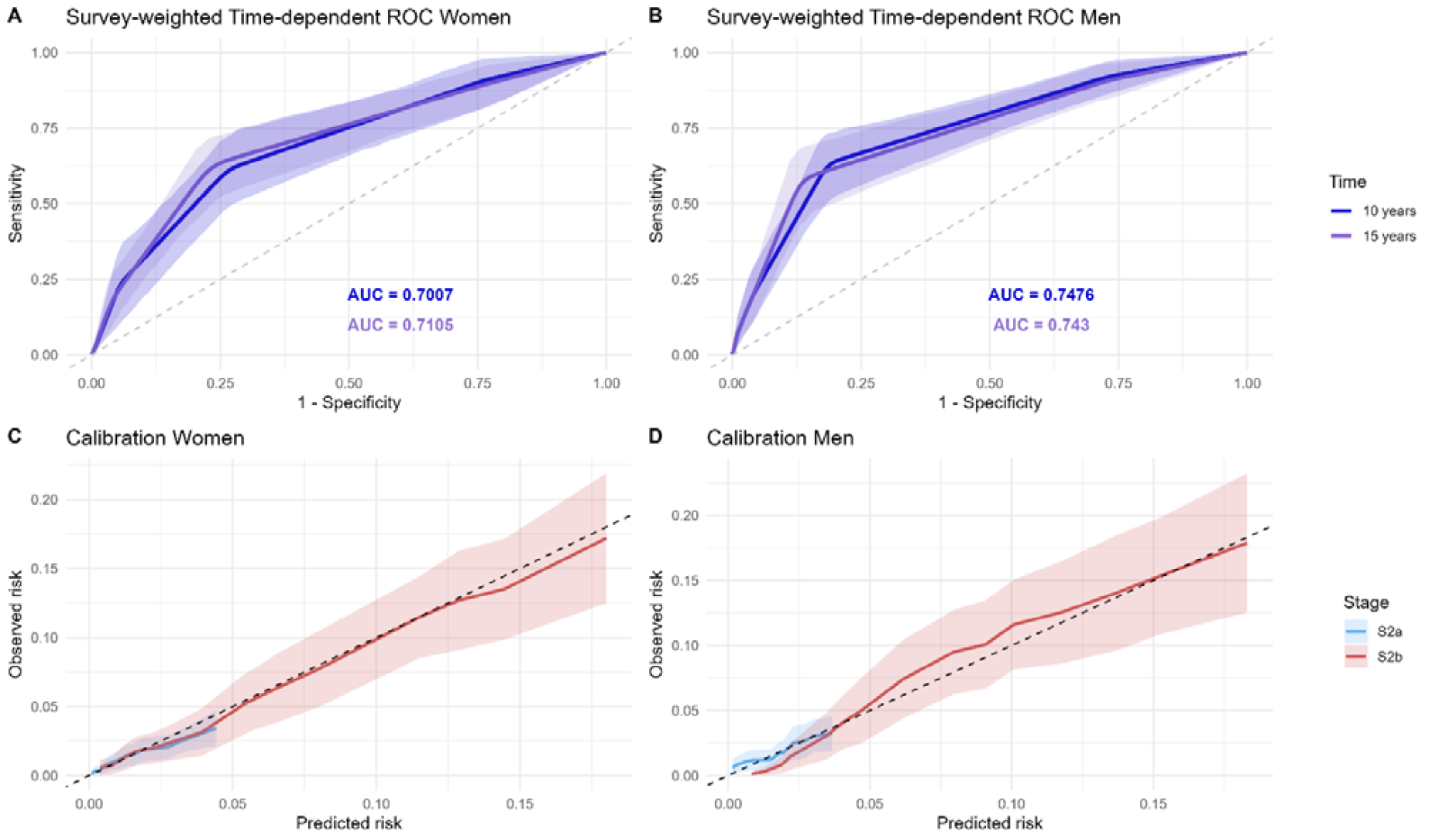
External Validation of the Sex-specific Risk-Stratification Model. Panels A and B present time-dependent ROC curves illustrating the discriminative performance of the sex-specific risk-stratification model for cardiovascular death in women (A) and men (B). Panels C and D show calibration plots for the same model in women (C) and men (D). *Abbreviations: AUC, area under the receiver operating characteristic curves; ROC, receiver operating characteristic*.

The 10-year cumulative incidences of cardiovascular death were 1.5% (95% CI 0.7-2.3) for women in stage 2a, 6.4% (95% CI 3.9-8.9) for women in stage 2b, 1.6% (95% CI 0.8-2.5) for men in stage 2a, and 10.1% (95% CI 6.7-13.5) for men in stage 2b (Figure 5). The absolute risk differences were 4.9% (95% CI 2.2-7.5) for women and 8.5% (95% CI 5.0-12.0) for men.

**Figure 5.**
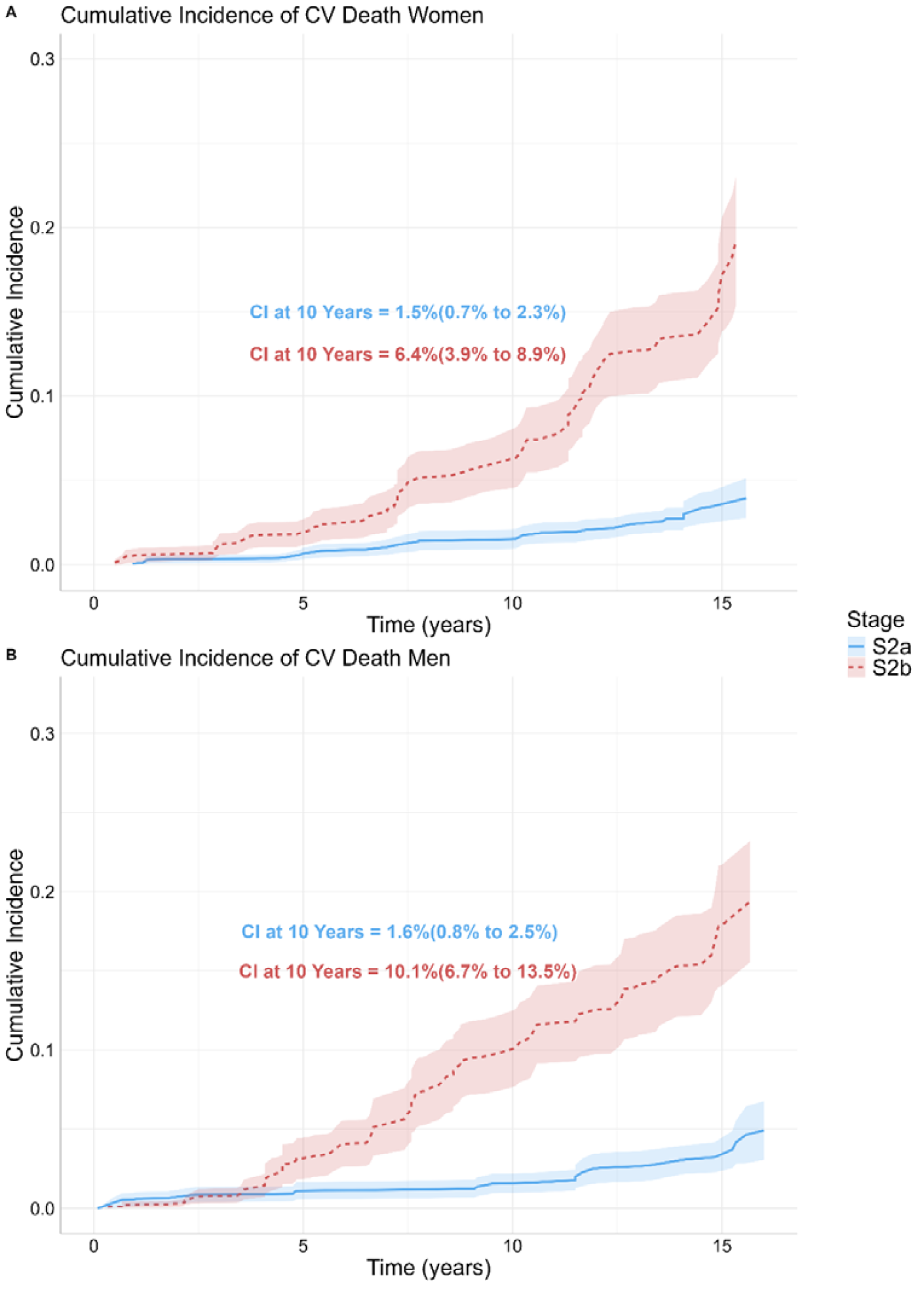
10-Year Cumulative Incidence of Cardiovascular Death among Participants with CKM Syndrome Stage 2a and Stage 2b in the Validation Cohort. Panels A and B depict weighted cumulative incidence curves for cardiovascular death in women and men, respectively, stratified by CKM syndrome stage 2a and 2b, with 95% confidence intervals. 10-year cumulative incidences are additionally presented as point estimates with 95% confidence intervals. *Abbreviations: CKM, cardiovascular-kidney-metabolic; CI, cumulative incidence*.

### Clinical Validation

A total of 223,224 women and 149,121 men in the TriNetX database met the criteria for CKM syndrome stage 2 and were included in the analysis (Supplemental Figure S5). The baseline characteristics were summarized in Table 2. The proportions of women and men with hypertension, MetS, hypertriglyceridemia, CKD, and diabetes were comparable. After applying the sex-specific risk-stratification model, the included participants were classified into four groups: women in stage 2a, women in stage 2b, men in stage 2a, and men in stage 2b.

**Table 2.** Baseline Characteristics of Participants in CKM syndrome Stage 2 identified in TriNetX database. Means are presented as estimated with standard deviations. Proportions were presented as estimates and 95% Confidence Intervals. Abbreviations: BMI, body mass index; CKD, chronic kidney disease; CKM, cardiovascular-kidneymetabolic; eGFR, estimated glomerular filtration rate; HbA1c, glycated hemoglobin; HDL, high density lipoprotein; HTG, hypertriglyceridemia; LDL, low density lipoprotein; MetS, metabolic syndrome; sysBP, systolic blood pressure; WC, waist circumference.

| Variable | TriNet X Cohort |  |
| --- | --- | --- |
|  | Women | Men |
| N | 169,580 | 156,340 |
| Age, year | 59.7 ± 14 | 59.1 ± 13.5 |
| White | 112,525 (50%) | 111,678 (58%) |
| Black | 69,008 (31%) | 43,376 (23%) |
| Asian | 12,309 (6%) | 10,372 (5%) |
| Hypertension | 145,133 (65%) | 125,807 (66%) |
| MetS | 144,751 (62%) | 131,086 (65%) |
| HTG | 5686 (3%) | 7351 (4%) |
| CKD | 29,133 (13%) | 28,582 (15%) |
| Diabetes | 151,863 (68%) | 130,732 (68%) |
| Smoking | 19,390 (9%) | 21,091 (11%) |
| Calcium channel blockers | 66,088 (28%) | 59,926 (30%) |
| ACE inhibitors/AT1-blocker | 66,539 (29%) | 70,808 (35%) |
| β Blockers | 69,814 (30%) | 59,926 (30%) |
| Statin | 118,216 (51%) | 112,093 (56%) |
| sysBP, mmHg | 132 ± 18.4 | 132 ± 17.8 |
| WC, cm | 104 ± 24.1 | 108 ± 25.6 |
| BMI, kg/m<sup>2</sup> | 33.2 ± 8.1 | 31.8 ± 6.9 |
| Total Cholesterol, mg/dl | 181 ± 44.7 | 166 ± 45.6 |
| HDL Cholesterol, mg/dl | 52.1 ± 16.7 | 43.4 ± 13.8 |
| LDL-Cholesterol, mg/dl | 100 ± 37.6 | 91.4 ± 36.4 |
| Triglyceride, mg/dl | 152 ± 117 | 169 ± 171 |
| eGFR, ml/m<sup>2</sup>/1.73m<sup>2</sup> | 81.4 ± 24.5 | 82.7 ± 23.5 |
| HbA1c, % | 7.1 ± 1.8 | 7.3 ± 1.9 |

During a mean follow-up of 3.8 years, 2280 MACE occurred among women in CKM syndrome stage 2a, and 8327 MACE in among women in stage 2b. Women in stage 2b experienced a more than three-fold higher cardiovascular risk than those in stage 2a (HR 3.29; 95% CI 3.14-3.44). The 10-year cumulative incidences of MACE differed markedly between the two groups: 7.0 % (95% CI 6.0-7.4) for stage 2a and 24.6% (95% CI 24.0-25.2) for stage 2b (Figure 6A). Among men, 4694 MACE were recorded in CKM syndrome stage⍰2a and 7920 MACE in stage⍰2b over a mean follow⍰up of 3.8⍰years. Compared with men in stage⍰2a, men in stage⍰2b had a more than two⍰fold higher cardiovascular risk (HR⍰2.32; 95%⍰CI⍰2.24-2.41). The 10⍰year cumulative incidence of MACE was substantially higher in men with stage⍰2b (30.8%; 95%⍰CI⍰30.0-31.5) than in those with stage⍰2a (13.1%; 95%⍰CI⍰12.7-13.6) (Figure 6B). These results demonstrate that the sex-specific risk-stratification model performs adequately for clinical use.

**Figure 6.**
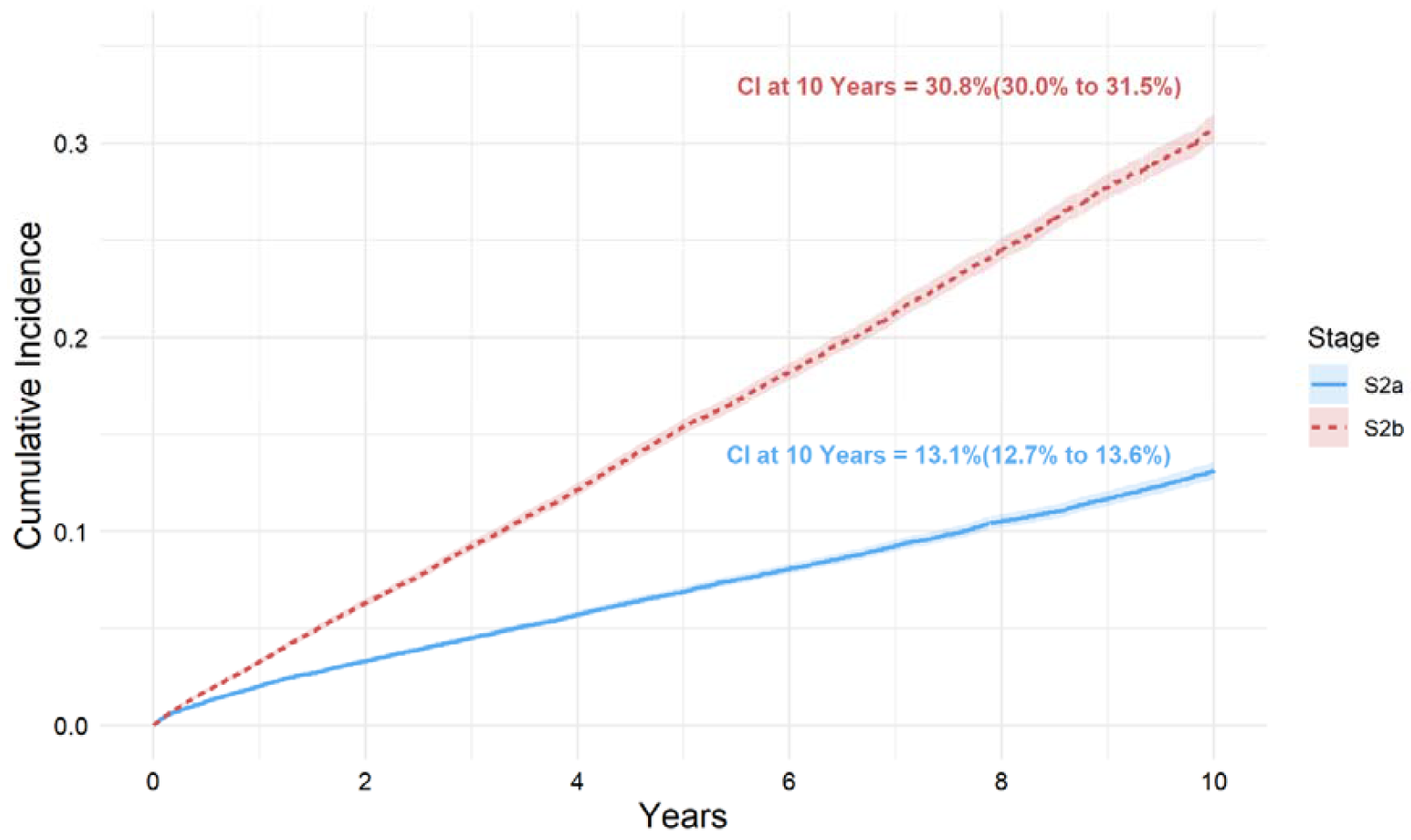
10-Year Cumulative Incidence of MACE among Participants with CKM Syndrome Stage 2a and Stage 2b identified in the TriNetX Database. Panels A and B depict cumulative incidence curves for MACE in women and men, respectively, stratified by CKM syndrome stage 2a and 2b, with 95% confidence intervals. 10-year cumulative incidences are additionally presented as point estimates with 95% confidence intervals. *Abbreviations: CKM, cardiovascular-kidney-metabolic; CI, cumulative incidence; MACE, major adverse cardiovascular events*.

## Discussion

Based on this nationally representative database of US adults, we developed and validated a sex-specific risk⍰stratification model that uses a simple algorithm to subdivide CKM syndrome stage⍰2 into two subgroups, which correlated with the degree of cardiovascular risk and demonstrated adequate stratification performance. In an external real-world clinical cohort, the model retained good discriminatory ability, effectively distinguishing individuals with higher cardiovascular risk from those at lower risk within CKD stage 2.

A bidirectional association between dysfunction of heart and kidney has been well-established. Metabolic abnormalities, such as diabetes and obesity, further shape this relationship and amplify cardiovascular risk. Conversely, kidney dysfunction also influences the link between metabolic abnormalities and CVD^26,27^. This interplay was emphasized in 2023 by AHA und summarized under the term CKM syndrome. A CKM syndrome staging system was also proposed with the aim of optimizing preventive strategies for earlier detection and timely intervention^1^. Prior studies have demonstrated that the CKM stages correlate well with cardiovascular death^3^. In particular, patients classified as CKM syndrome stage⍰2, who already exhibit organ damage such as chronic kidney disease or metabolic syndrome but have not yet developed CVD, constitute roughly half of the US adult population. This group experiences an approximately two⍰fold higher risk of cardiovascular death compared with individuals in stage⍰0, however, the magnitude of the risk is markedly modified by age and sex^3,8^. As a result, a subset of stage 2 patients with substantially elevated cardiovascular risk may be overlooked in primary⍰care settings and consequently receive suboptimal treatment under the current staging system. This underscores the need to further refine CKM syndrome stage⍰2 by a straightforward algorithm that can be easily applied by primary⍰care clinicians. By implementing our risk⍰stratification model in clinical practice, we expect that the current gap can be closed.

Accumulating evidence shows how sex influences cardiovascular health through its interaction with multiple aspects, such as health⍰behavior patterns, hormonal and other biological differences, and psychosocial stress^28–30^. To highlight sex related aspects, we analyzed women and men separately to develop a sex⍰specific model. Despite differing sex-specific associations of diverse variables, diabetes and CKD were included in the final model for both women and men in CKM syndrome stage 2, while a more pronounced effect of them in women was observed in both LASSO selection and Cox⍰regression. As the most common type of diabetes, type 2 diabetes mellitus accounts for approximately 90% of all cases and its prevalence is similar in women and men^31,32^. Diabetes is a major cardiovascular risk factor for both sexes; however, sex⍰specific differences have been observed. Diabetic women appear to experience a markedly greater adverse impact of diabetes, with a steeper rise in mortality compared with diabetic men^33^. Similar to diabetes, CKD is also recognized as a well-established amplifier of cardiovascular risk^34^ and exhibits sex⍰specific differences with cardiovascular events as outcome^29,35–37^. Notably, the deleterious cardiovascular effects of CKD are evident across all levels of eGFR and albuminuria^38^. Moreover, the excess cardiovascular risk associated with CKD is greater in women than in men, an effect that is consistent across strata of eGFR, age, and blood pressure^29^. The most notable sex⍰specific differences were the stronger link between hypertension and cardiovascular risk in women and the greater relevance of smoking in men. Current hypertension guidelines are identical for both sexes, yet important sex⍰specific differences have been reported. Across the life course, women experience a steeper rise in blood pressure already beginning in their 20s^39^. Moreover, an elevated cardiovascular risk is observed in women at “healthy” systolic blood pressure (BP) levels, while men only show increased risk when systolic BP reaches the abnormal range (≥⍰130⍰mm⍰Hg)^40–42^. Finally, obesity and dyslipidemia appear to play only a minor role in individuals without established CVD.

In clinical practice, the present study demonstrates that CKM syndrome stage 2b confers a markedly elevated cardiovascular risk in both sexes, underscoring the need for tailored preventive and therapeutic strategies. An intensive lifestyle⍰intervention program should be instituted as a fundamental component of care for this population. Based on current evidence, intensive BP control should be implemented because it confers a net cardiovascular benefit^43–46^, and a lower systolic BP target may be appropriate for women at CKM syndrome stage⍰2b. In addition, recent large-scale clinical trials have demonstrated promising protective effects on cardiovascular health of sodium-glucose cotransporter-2 inhibitors, glucose-like peptide-1 receptor agonists, and finerenone^47–56^. Patients classified as CKM⍰syndrome stage⍰2b are frequently obese and often have CKD and/or diabetes. Consequently, they are likely to benefit most from a broad and equitable implementation of these therapies in primary⍰care settings.

Strengths of this study include a nationally representative sample with a long follow⍰up period for model development, a sequential covariate selection strategy, and external validation of model performance also using real⍰world clinical data. Nevertheless, several limitations remain. First, CVD in NHANES was identified solely by self-report during house interview. Definitive diagnostic modalities such as echocardiography, myocardial scintigraphy, cardiac magnetic⍰resonance imaging, or coronary angiography were not available in NHANES. Consequently, some participants may have been misclassified. Second, Asian participants were not classified and oversampled until 2011. This subgroup population may be under-represented in our study. Therefore, the risk-stratification model should be applied to Asian patients with caution. Furthermore, the risk-stratification model could be improved with incorporating incident major cardiac events as outcomes. However, the longitudinal data of NHANES captured only cardiovascular death. The TriNetX database lacks cause-specific mortality information, which prompted us to choose a different outcome in this cohort. Finally, neither NHANES nor TriNetX distinguishes biological sex assigned at birth from gender identity. Consequently, the staging of individuals whose gender identity does not align with their sex assigned at birth is uncertain, and applying our risk⍰stratification model to this subgroup may be problematic. Although the model is defined on the basis of biological sex, it cannot disentangle gender⍰specific differences.

In conclusion, we have developed and validated a risk⍰stratification model that uses a simple algorithm to subdivide CKM syndrome stage⍰2 into two distinct subgroups. The model is intended as an additional tool for more tailored and precise patient management in primary-care settings, with the goal of preventing progression to advanced stages. Individuals classified as CKM syndrome stage⍰2b may particularly benefit from early⍰intervention strategies guided by this approach.

## Abbreviations

AHA: American Heart Association
AUC: Area under the receiver operating characteristic curve
CI: Confidence intervall
CKD: Chronic kidney disease
CKM: syndrome Cardiovascular-kidney-metabolic syndrome
CVD: Cardiovascular disease
eGFR: Estimated glomerular filtration
HR: Hazard ratio
LASSO: Least absolute shrinkage and selection operator
MACE: Major adverse cardiac events
MetS: Metabolic syndrome
NHANES: National Health and Nutrition Examination Survey
UACR: Urinary albumin to creatinine ratio

## Acknowledgments

No other individuals contributed to this study substantially.

## Author contributions

Z. Tian participated in research concept, research design, data acquisition, statistical analysis and drafting of the manuscript.

M. Sternberg participated in data acquisition and statistical analysis.

J. Willerding and K. M. Schmidt-Ott participated in reviewing the contents.

A. Melk and B. M. W. Schmidt participated in research concept, research design, statistical analysis, supervision and reviewing the contents.

## Data Sharing Statement

NHANES data are available from the NHANES web page. The data supporting this study’s findings are available from TriNetX, LLC, but restrictions apply to their availability. The data were accessed under license and cannot be publicly shared. Accredited researchers may obtain access through a data use agreement with TriNetX, LLC, which may involve licensing fees. The R code of the analysis is available from the authors on reasonable request.

